# Early antibiotics, mode of delivery and social-emotional problems in childhood: The FinnBrain Birth Cohort Study

**DOI:** 10.64898/2025.12.22.25342801

**Authors:** Jenni Korteniemi, Anna-Katariina Aatsinki, Laura Perasto, Hasse Karlsson, Katja Tervahartiala, Linnea Karlsson

## Abstract

**Objective:** Both early life antibiotics and the mode of delivery have been associated with multiple somatic health outcomes but less is known about their possible psychosocial outcomes. The aim of this study was to study whether early antibiotic treatments and mode of delivery are associated with social and emotional problems in children. Studies have shown that modulating gut microbiota may have sex-specific effects on brain-related and behavioral outcomes, therefore we expected that these exposures would associate with child outcomes in a sex-specific manner.

**Methods:** Subjects (n=1766) were recruited from the FinnBrain Birth Cohort study in Finland. Register data on mode of delivery, mother’s prenatal antibiotics and child antibiotics was combined with questionnaire data to investigate the associations between the mode of delivery and antibiotic treatments on child social and emotional problems in cohort population. Social and emotional problems were measured with Brief Infant-Toddler Social and Emotional Assessment at 2 years and Strengths and Difficulties Questionnaire at 5 years. The associations were analyzed using non-parametric Wilcoxon rank sum test or Kruskal-Wallis test.

**Results:** Neither the mode of delivery nor early life antibiotic treatments were associated with social and emotional problems in children. Sex-stratified analyses yielded similar results. The combined exposure of mode of delivery and early antibiotic use yielded nonsignificant associations.

**Conclusions:** Our data did not support the hypothesis of the associations between birth mode and early antibiotics and child social-emotional problems. Longitudinal studies focusing on clinically severe phenotypes and specific risk groups might gain better understanding of this topic.

## Introduction

There is growing evidence for early life exposures to be involved in the development of mental disorders^1–3^. One potential pathway for this association has been thought to be the microbiota-gut-brain-axis^4^ with the first 2 years of life being recognized as a critical window for the maturing of gut microbiota (GM)^5–8^. In early childhood, GM is affected especially by mode of delivery, antibiotics and diet^9–12^. While there is clear evidence for systemic effect of antibiotics and mode of delivery on somatic health outcomes such as asthma and obesity^13,14^, less is known about possible social-emotional outcomes.

There are several studies that have investigated the association between caesarean section (C-section) and later emotional and behavioral outcomes, with most existing studies done in age groups including both children and adolescents and even young adults^15–17^. Studies that have focused on social-emotional development in younger children are rather heterogenous, with child age at phenotype assessment varying from 9 months up to 17 years^18,19^ and some classifying delivery mode in two (C-section and vaginal delivery) and others in multiple categories, such as assisted and unassisted vaginal delivery and planned and emergency C-section^20,21^. However, there is some evidence indicating that C-section might be associated with behavioral difficulties and delays in motor and social skill development in children from the age of three onwards^18,22,23^.

Research on early antibiotics and later psychiatric outcomes has mostly included animal studies, with preliminary results indicating that early life antibiotic treatments might result in altered behavior via changes in GM composition, blood-brain barrier integrity and neuromodulators^24–26^. Moreover, there are several human studies where antibiotics during pregnancy and first two years of life have been associated with higher risk for neuropsychiatric and other mental disorders^27–30^. Existing studies have largely focused on diagnosed neuropsychiatric and affective disorders as outcomes. There is less knowledge available on whether early exposure to antibiotics is associated with social-emotional developmental outcomes in early childhood, potential precursors of later (neuro)psychiatric disorders.

In this study we aimed to examine whether mode of delivery and early antibiotic treatments during pregnancy, around birth and first two years of life associate with later social and emotional problems in children and how this connection is moderated by child’s biological sex assigned at birth. In addition, the frequency and timing of antibiotic exposure as well as combined effects of both C-section and antibiotics exposure were explored. We combined register data on antibiotic purchases and perinatal factors with questionnaire data investigating child social and emotional problems at 2 and 5 years in FinnBrain Birth Cohort population. Multiple studies have shown that modulating GM may have sex-specific effects on brain-related and behavioral outcomes, therefore we expected that these early life exposures associate with child outcomes in sex-specific manner. Given that behavioral profiles in males are often reported to be more affected by early life microbiome modulation, we hypothesize that boys show stronger associations.

## Methods

### Study population

Subjects were recruited from the FinnBrain Birth Cohort study in South-Western Finland and Åland Islands (mothers n=3808, children n=3898)^31^. Parents were recruited during the first trimester ultrasound visit at gestational week 12. Parents sufficiently fluent in Finnish or Swedish, and with normal first trimester ultrasound screening result were included in the cohort. The children were born between 2012 and 2015. In this study, multiple births and unknown modes of delivery were excluded. Subjects with both relevant register data and questionnaire data available were included in the final analyses (Figure 1).

**Figure 1.**
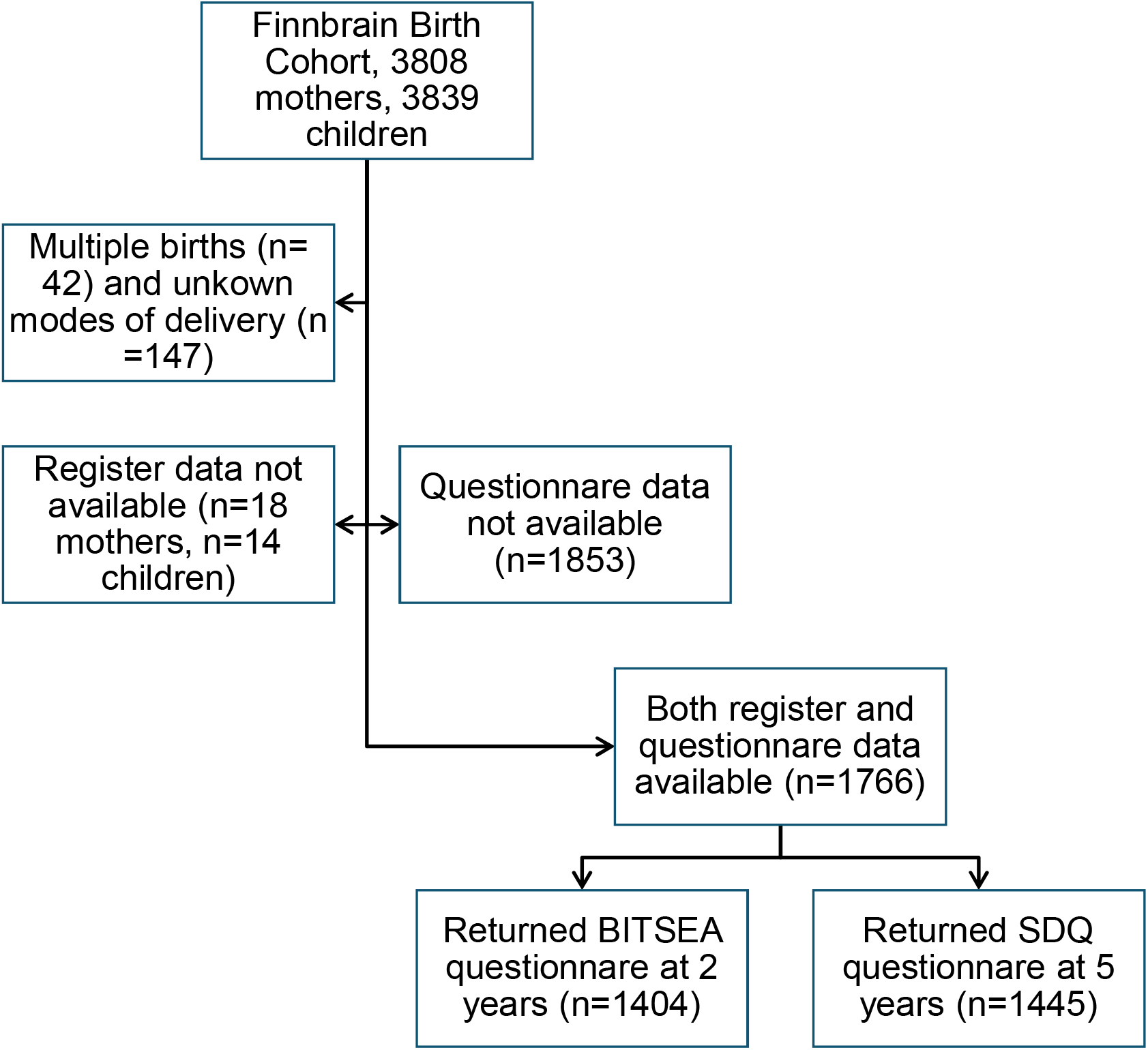
Flow chart on participant selection of this study.

### Exposure

Data on medication purchases including mother’s prenatal antibiotics and child antibiotics from the first two years were acquired from Finnish Social Insurance Institution registers that contain all purchases of prescribed medications. In Finland purchasing antibiotics is only possible with a doctor’s prescription. We included all purchases of oral antibiotics. Information about child’s perinatal antibiotics up to 7 days postpartum was received from hospital records. All Finnish residents have unique personal identification numbers which makes combining data from multiple registers possible. Exposure to antibiotics was categorized based on the number of treatments: 0, 1 to 3, and 4 or more courses of antibiotic treatment. The timing of antibiotics was categorized in three groups: antibiotics during pregnancy, around birth and the first two years of life.

Information on the mode of delivery was received from hospital records. The mode of delivery was categorized in four groups based on previous literature: unassisted vaginal birth, assisted vaginal birth, planned C-section and emergency C-section. Assisted vaginal birth included the induction of birth and if equipment was used during the delivery. We also did the analyses using just two categories: all vaginal births and all C-sections.

Since we wanted to explore the association between mode of delivery and antibiotics together with the outcome, we then divided the subjects in four groups: vaginal delivery and no antibiotic treatments, vaginal delivery and antibiotic treatments, C-section but no antibiotic treatments and finally, C-section and antibiotic treatments. The analyses were done three times based on the timing of antibiotics (during pregnancy, perinatal, first 2 years of life).

### Measures

To evaluate the possible psychosocial outcomes, child social-emotional problems was measured with Brief Infant-Toddler Social and Emotional Assessment (BITSEA) at 2 years and Strengths and Difficulties Questionnaire (SDQ) at 5 years. Both questionnaires are standardized and internationally widely used and were filled in by mothers^32,33^. The BITSEA contains 42 items (rated from 0=Not true to 2=Very true) developed for screening socio-emotional and behavioral problems and competence among toddlers aged 12-36 months. The questionnaire includes factors measuring internalizing and externalizing problems, dysregulation, autism spectrum disorders, and rear clinically relevant red flag items. In this study, only the Total Problem scale was used which is the sum score of problem measures. Cronbach’s alpha for BITSEA Total problem score was 0,7 in our sample. The SDQ is a widely used instrument for measuring psychosocial problems and competence in children ages 3-16 years. It consists of 25 items (rated from 0=Not true to 2=Certainly true) divided in five scales: Emotional symptoms, Conduct problems, Hyperactivity-inattention problems, Peer relationship problems, and Prosocial behavior. In our analyses, only the Total Problem Score, which is sum score of problem scales, was used. Cronbach’s alpha for SDQ Total Problem score was 0,8 in our sample.

### Background factors

*A priori* identified confounders in this study were child’s weight for gestational age, the 5-minutes Apgar-score (>7 and <7 points as categorical variable)^34^, mother’s body mass index (BMI) and age, parity, breastfeeding status, socioeconomic status, smoking during pregnancy, mother’s depressive symptoms during pregnancy and mother’s any somatic disorders and medications (supplemental materials, Figure 1).

Participant data were obtained from cohort and hospital records. Maternal depressive symptoms were assessed using the Edinburgh Postnatal Depression Scale^35^, while maternal pre-pregnancy BMI, parity, multiple births, birth weight category (small, appropriate, large for gestational age), maternal diagnoses and medications and Apgar-score were retrieved from hospital records. Child sex was defined as biological sex assigned at birth.

### Statistical analyses

First, the associations between categorical delivery mode and child’s continuous Total Problem scores (BITSEA and SDQ) were analyzed using nonparametric Wilcoxon rank sum test or Kruskall-Wallis test. With binary delivery mode (C-section and vaginal) Wilcoxon rank sum was used and with four class delivery mode (assisted vaginal, vaginal, planned C-section, emergency C-section) Kruskall-Wallis was used.

Next, the associations between antibiotic exposure and child’s continuous total scores were analyzed in the same manner. Three different antibiotics variables were studied: 1) binary variable indicating whether mother had gotten antibiotics during pregnancy, 2) binary variable indicating whether child had gotten antibiotics during delivery and 3) three class variable indicating how many antibiotics the child had had during the first 2 years of life (0, 1-3, 4 or more).

Lastly, binary delivery mode and antibiotic variables were combined. Associations with this combination variable and total scores were examined using Kruskall-Wallis test. In the case ofstatistically significant findings, a regression model including *a priori* identified potential confounders would have been constructed. P-values (two-tailed) < 0.05 were considered statistically significant.

## Results

1766 subjects with relevant data were included in this study, of which 814 (46.1%) were girls and 952 (53.9%) were boys. In all, 1404 mothers returned BITSEA questionnaire at the child mean age of 2.05 years (SD 0.05 years) and 1592 mothers returned SDQ questionnaire at the child mean age of 5.20 years (SD 0.24 years). Attrition analysis showed that respondents were on average older, more highly educated and primiparous than non-respondents. There was no statistically significant difference in birth mode or register data availability between respondents and non-respondents (supplemental materials, Table 1). In this study sample, 1462 (82.8%) of children overall were born with vaginal delivery, compared to 304 (17.2%) born with C-section. 1362 subjects (77.1%) had antibiotic purchases either before pregnancy, around delivery or the first 2 years of life (Tables 1–3).

**Table 1.** Descriptive information of study sample.

|  | Boys<br>(N=952) | Girls<br>(N=814) | Overall<br>(N=1766) |
| --- | --- | --- | --- |
| **Size for GA\*** |  |  |  |
| SGA, n (%) | 30 (3.2) | 15 (1.8) | 45 (2.5) |
| AGA, n (%) | 902 (94.7) | 776 (95.3) | 1678 (95.0) |
| LGA, n (%) | 20 (2.1) | 23 (2.8) | 43 (2.4) |
| Missing, n (%) | 0 (0) | 0 (0) | 0 (0) |
| **Apgar at 5 min** |  |  |  |
| Mean (SD) | 9.03 (0.82) | 9.06 (0.85) | 9.05 (0.83) |
| Median [Min, Max] | 9.00 [3.0, 10.00] | 9.00 [3.00, 10.00] | 9.00 [3.00, 10.00] |
| Missing, n (%) | 5 (0.5) | 2 (0.2) | 7 (0.4) |
| **Mother's age at delivery** |  |  |  |
| Mean (SD) | 30.89 (4.26) | 30.97 (4.40) | 30.93 (4.32) |
| Median [Min, Max] | 31.00 [17.00, 42.00] | 31.00 [18.00, 45.00] | 31.00 [17.00, 45.00] |
| Missing, n (%) | 0 (0) | 0 (0) | 0 (0) |
| **Mother's prenatal BMI** |  |  |  |
| Mean (SD) | 24.37 (4.63) | 24.68 (4.99) | 24.51 (4.80) |
| Median [Min, Max] | 23.30 [16.56, 47.88] | 23.53 [16.90, 60.61] | 23.39 [16.56, 60.61] |
| Missing, n (%) | 15 (1.6) | 13 (1.6) | 28 (1.6) |
| **Smoking during pregnancy** |  |  |  |
| Yes, n (%) | 123 (12.9) | 97 (11.9) | 220 (12.5) |
| No, n (%) | 829 (87.1) | 717 (88.1) | 1546 (87.5) |
| Missing, n (%) | 0 (0) | 0 (0) | 0 (0) |
| **Parity** |  |  |  |
| Primipara, n (%) | 501 (52.6) | 429 (52.7) | 930 (52.7) |
| Multipara, n (%) | 451 (47.4) | 385 (47.3) | 836 (47.3) |
| Missing, n (%) | 0 (0) | 0 (0) | 0 (0) |
| **Breastfeeding** |  |  |  |
| Fully breastfed, n (%) | 570 (64.3) | 501 (66.4) | 1071 (65.3) |
| Partial, n (%) | 300 (33.8) | 241 (32.0) | 541 (33.0) |
| None, n (%) | 17 (1.9) | 12 (1.6) | 29 (1.8) |
| Missing, n (%) | 65 (6.8) | 60 (7.4) | 125 (7.1) |
| **Maternal years of education** |  |  |  |
| < 12 years, n (%) | 288 (31.5) | 223 (28.7) | 511 (30.2) |
| 12–15 years, n (%) | 266 (29.1) | 239 (30.8) | 505 (29.9) |
| > 15 years, n (%) | 360 (39.4) | 314 (40.5) | 674 (39.9) |
| Missing, n (%) | 38 (4.0) | 38 (4.7) | 76 (4.3) |
| **Maternal somatic disorders** |  |  |  |
| No, n (%) | 747 (78.5) | 660 (81.1) | 1407 (79.7) |
| Yes, n (%) | 205 (21.5) | 154 (18.9) | 359 (20.3) |
| Missing, n (%) | 0 (0) | 0 (0) | 0 (0) |

\* GA= gestational age
SGA= Small for GA
AGA= Appropriate for GA
LGA= Large for GA
BMI= Body mass index

There was no significant difference in child BITSEA problem score or SDQ total sum neither by the mode of delivery nor by maternal or child antibiotics exposure at any time point (Table 4, supplemental materials Table 2). Similarly, cumulative number of antibiotic purchases was not related to BITSEA problem score or SDQ total sum (Table 4, supplemental materials Table 2). Combination of C-section and antibiotic exposure was not related to BITSEA or SDQ, regardless of antibiotic timing (Table 4, supplemental materials Table 2). The results were similar in both sexes and there were no significant differences between groups (Table 4, supplemental materials Table 2 and Figures 2-3).

**Table 2.**
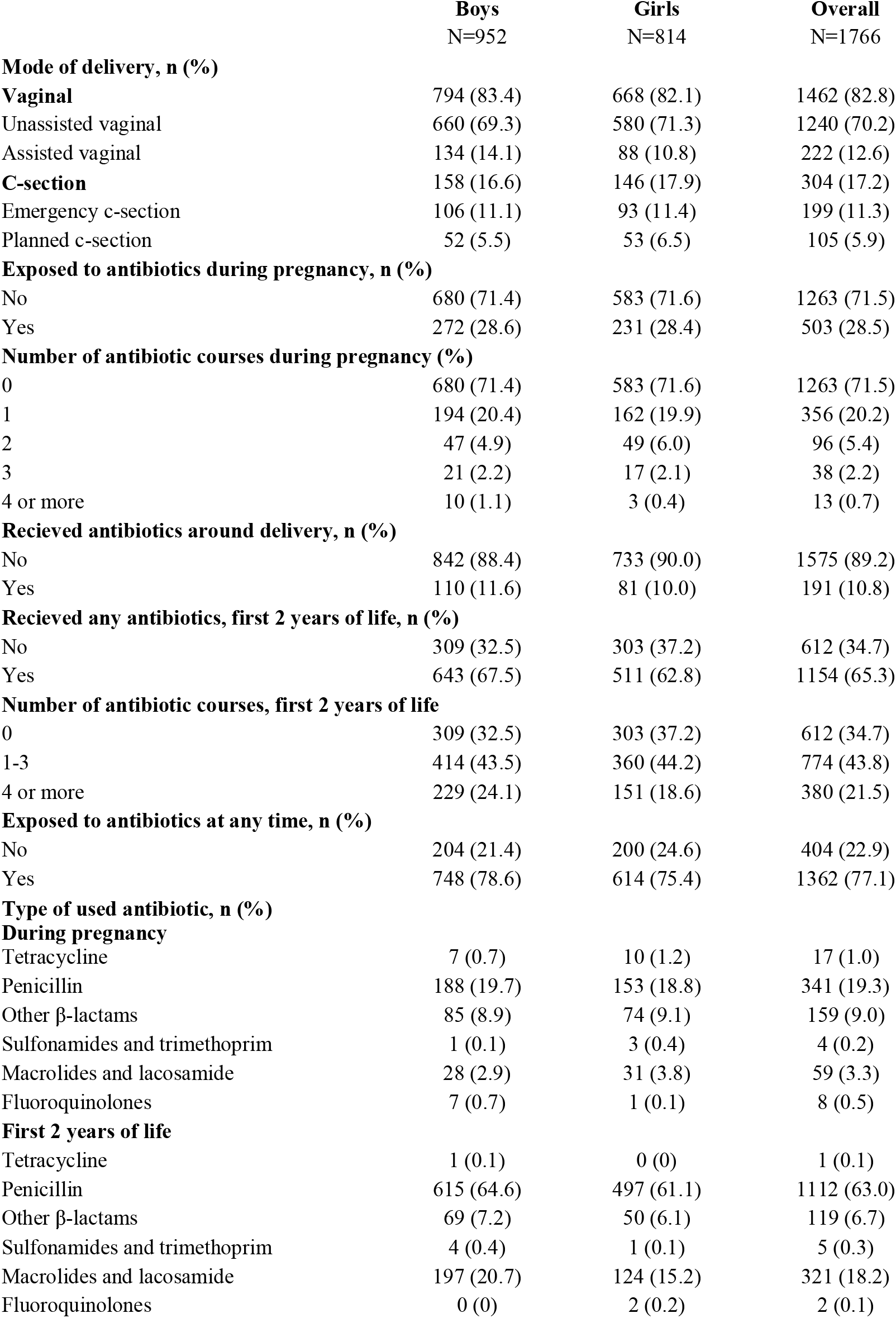
Information on mode of delivery and antibiotic treatment rates in study sample.

|  | **Boys**<br>N=952 | **Girls**<br>N=814 | **Overall**<br>N=1766 |
| --- | --- | --- | --- |
| **Mode of delivery, n (%)** |  |  |  |
| **Vaginal** | 794 (83.4) | 668 (82.1) | 1462 (82.8) |
| Unassisted vaginal | 660 (69.3) | 580 (71.3) | 1240 (70.2) |
| Assisted vaginal | 134 (14.1) | 88 (10.8) | 222 (12.6) |
| **C-section** | 158 (16.6) | 146 (17.9) | 304 (17.2) |
| Emergency c-section | 106 (11.1) | 93 (11.4) | 199 (11.3) |
| Planned c-section | 52 (5.5) | 53 (6.5) | 105 (5.9) |
| **Exposed to antibiotics during pregnancy, n (%)** |  |  |  |
| No | 680 (71.4) | 583 (71.6) | 1263 (71.5) |
| Yes | 272 (28.6) | 231 (28.4) | 503 (28.5) |
| **Number of antibiotic courses during pregnancy (%)** |  |  |  |
| 0 | 680 (71.4) | 583 (71.6) | 1263 (71.5) |
| 1 | 194 (20.4) | 162 (19.9) | 356 (20.2) |
| 2 | 47 (4.9) | 49 (6.0) | 96 (5.4) |
| 3 | 21 (2.2) | 17 (2.1) | 38 (2.2) |
| 4 or more | 10 (1.1) | 3 (0.4) | 13 (0.7) |
| **Received antibiotics around delivery, n (%)** |  |  |  |
| No | 842 (88.4) | 733 (90.0) | 1575 (89.2) |
| Yes | 110 (11.6) | 81 (10.0) | 191 (10.8) |
| **Received any antibiotics, first 2 years of life, n (%)** |  |  |  |
| No | 309 (32.5) | 303 (37.2) | 612 (34.7) |
| Yes | 643 (67.5) | 511 (62.8) | 1154 (65.3) |
| **Number of antibiotic courses, first 2 years of life** |  |  |  |
| 0 | 309 (32.5) | 303 (37.2) | 612 (34.7) |
| 1-3 | 414 (43.5) | 360 (44.2) | 774 (43.8) |
| 4 or more | 229 (24.1) | 151 (18.6) | 380 (21.5) |
| **Exposed to antibiotics at any time, n (%)** |  |  |  |
| No | 204 (21.4) | 200 (24.6) | 404 (22.9) |
| Yes | 748 (78.6) | 614 (75.4) | 1362 (77.1) |
| **Type of used antibiotic, n (%)** |  |  |  |
| **During pregnancy** |  |  |  |
| Tetracycline | 7 (0.7) | 10 (1.2) | 17 (1.0) |
| Penicillin | 188 (19.7) | 153 (18.8) | 341 (19.3) |
| Other β-lactams | 85 (8.9) | 74 (9.1) | 159 (9.0) |
| Sulfonamides and trimethoprim | 1 (0.1) | 3 (0.4) | 4 (0.2) |
| Macrolides and lacosamide | 28 (2.9) | 31 (3.8) | 59 (3.3) |
| Fluoroquinolones | 7 (0.7) | 1 (0.1) | 8 (0.5) |
| **First 2 years of life** |  |  |  |
| Tetracycline | 1 (0.1) | 0 (0) | 1 (0.1) |
| Penicillin | 615 (64.6) | 497 (61.1) | 1112 (63.0) |
| Other β-lactams | 69 (7.2) | 50 (6.1) | 119 (6.7) |
| Sulfonamides and trimethoprim | 4 (0.4) | 1 (0.1) | 5 (0.3) |
| Macrolides and lacosamide | 197 (20.7) | 124 (15.2) | 321 (18.2) |
| Fluoroquinolones | 0 (0) | 2 (0.2) | 2 (0.1) |

**Table 3.**
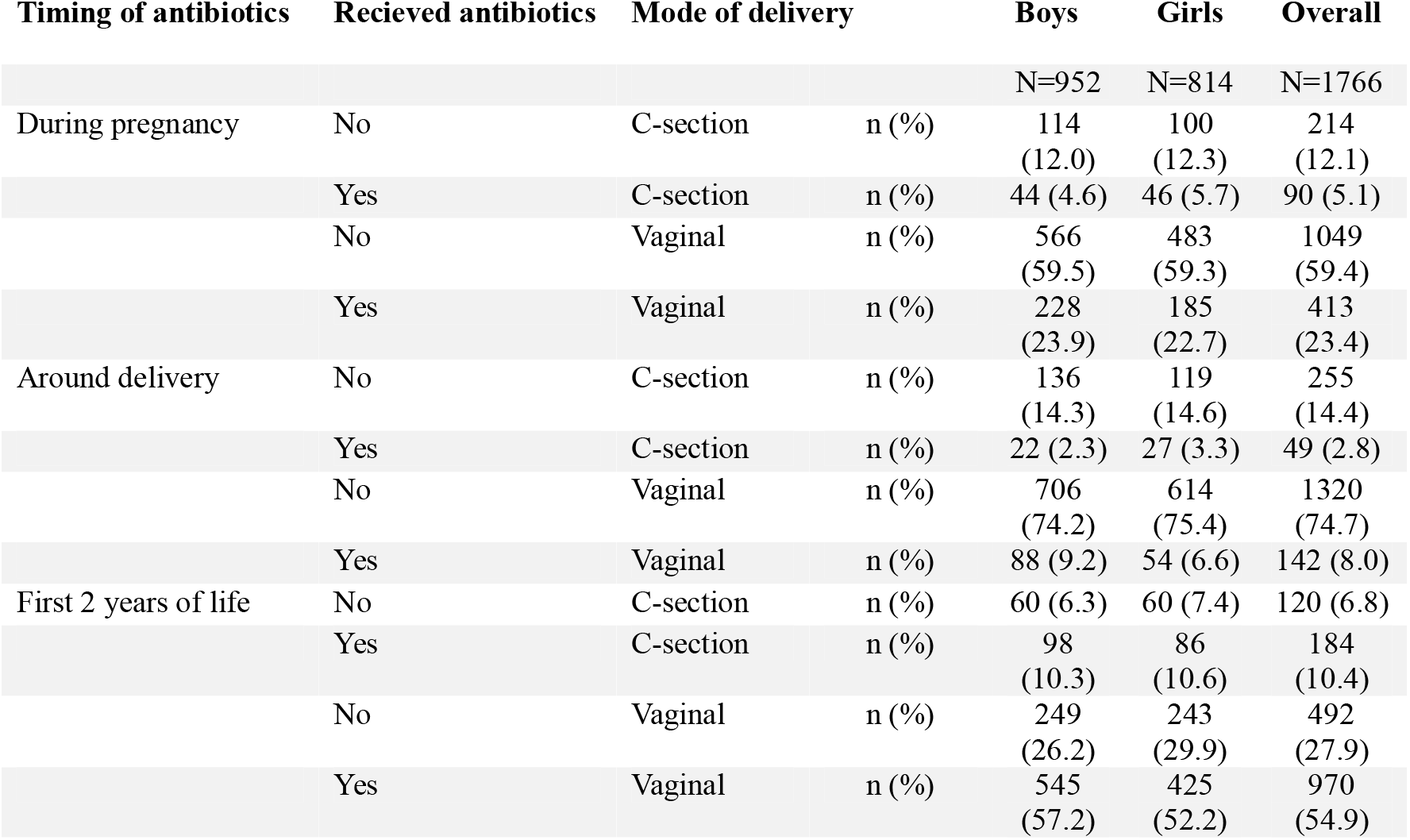
Information on antibiotic treatment rates based on mode of delivery in study sample.

**Table 4.** Used statistical tests and p-values when comparing different exposures with SDQ and BITSEA scores in both sexes and when stratified by sex.

| Questionnaire score used | Exposure | n | Test used | p-value |
| --- | --- | --- | --- | --- |
| Bitsea Problem | AB* prenatal | (AB / No AB) | wilcoxon rank sum | .826 |
|  | Girl | 238/596 |  | .281 |
|  | Boy | 278/698 |  | .463 |
| Bitsea Problem | AB perinatal | (AB / No AB) | wilcoxon rank sum | .892 |
|  | Girl | 86/748 |  | .0789 |
|  | Boy | 113/863 |  | .196 |
| Bitsea Problem | AB courses, first 2 yrs of life | (0/1-3/4 or more) | kruskal-wallis | .0809 |
|  | Girl | 308/373/153 |  | .109 |
|  | Boy | 316/422/238 |  | .665 |
| SDQ sum | AB prenatal |  | wilcoxon rank sum | .803 |
|  | Girl |  |  | .758 |
|  | Boy |  |  | .521 |
| SDQ sum | AB perinatal |  | wilcoxon rank sum | .0948 |
|  | Girl |  |  | .571 |
|  | Boy |  |  | .152 |
| SDQ sum | AB courses, first 2 yrs of life |  | kruskal-wallis | .668 |
|  | Girl |  |  | .606 |
|  | Boy |  |  | .423 |
| Bitsea Problem | Delivery mode, 4 cat* | AVD/ECS/PCS/UVD* | kruskal-wallis | .413 |
| Girl |  | 89/94/54/597 |  | .793 |
| Boy |  | 137/109/54/676 |  | .650 |
| SDQ sum | Delivery mode, 4 cat |  | kruskal-wallis | .0593 |
| Girl |  |  |  | .318 |
| Boy |  |  |  | .345 |
| Bitsea Problem | Delivery mode, 2 cat | CS/VD | wilcoxon rank sum | .435 |
| Girl |  | 148/686 |  | .620 |
| Boy |  | 163/813 |  | .663 |
| SDQ sum | Delivery mode, 2 cat |  | wilcoxon rank sum | .439 |
| Girl |  |  |  | .356 |
| Boy |  |  |  | .821 |
| Bitsea Problem | AB (prenatal) + DM* combination | CS+noAB/CS+AB/VD+noAB/VD+AB | kruskal-wallis | .229 |
| Girl |  | 102/46/494/192 |  | .137 |
| Boy |  | 11944/579/234 |  | .690 |
| SDQ sum | AB (prenatal) + DM combination |  | kruskal-wallis | .870 |
| Girl |  |  |  | .166 |
| Boy |  |  |  | .483 |
| Bitsea Problem | AB (perinatal) + DM combination | CS+noAB/CS+AB/VD+noAB/VD+AB | kruskal-wallis | .573 |
| Girl |  | 120/28/628/58 |  | .162 |
| Boy |  | 141/22/722/91 |  | .575 |
| SDQ sum | AB (perinatal) + DM combination |  | kruskal-wallis | .299 |
| Girl |  |  |  | .689 |
| Boy |  |  |  | .533 |
| Bitsea Problem | AB (first 2 yrs) + DM combination | CS+noAB/CS+AB/VD+noAB/VD+AB | kruskal-wallis | .701 |
| Girl |  | 61/87/247/439 |  | .905 |
| Boy |  | 62/101/254/559 |  | .910 |
| SDQ sum | AB (first 2 yrs) + DM combination |  | kruskal-wallis | .649 |
| Girl |  |  |  | .166 |
| Boy |  |  |  | .500 |

\*AB = Recieved antibiotics
no AB = did not recieve antibiotics
Cat= categories
AVD= Assisted vaginal delivery
ECS = Emergency caesarean section
PCS= Planned caesarean section (elective caesarean section)
UVD = unassisted vaginal delivery
CS = all caesarean secrions
VD= all vaginal deliveries
DM = Delivery mode

## Discussion

We investigated the relations between prenatal and early life antibiotics and delivery mode and child social-emotional problems in population-based cohort that included 1766 participants. Our results did not imply any significant association between mode of delivery, early life antibiotic treatments, and later social-emotional problems in young children. The results were similar in girls and boys, and neither the timing (i.e. during pregnancy compared to infancy) nor the cumulative number of antibiotics altered the observation.

Our results regarding delivery mode and child social-emotional problems are consistent with previous cohort studies by Khalaf et al., Takács et al. and Maher et al.^18,19,23^, all of them including SDQ scores in similar time points as we did (9 months, 3, 4 and 5 years, but also adolescents). Takács et al. did not distinguish elective and emergency C-section due to small sample size, and in the studies by Khalaf et al. and Maher et al. mode of delivery was self-reported, not from register data. There are no comparable studies using BITSEA scores. However, our findings are contrary to previous register-based cohort studies using psychiatric diagnoses as outcomes and including older children and adolescents^3,15–17^. In a recent systematic review and meta-analysis by Green et al. combining 30 studies and over 7 million participants aged 2-18 years, there was a weak association between early antibiotics and later neurodevelopmental outcomes, especially in autism spectrum disorders (ASD) and attention-deficit hyperactivity disorder (ADHD)^36^. However, association was not found in sibling-controlled subgroup analyses indicating that the link is likely confounded by other environmental or genetic factors. Most studies included in the meta-analysis were retrospective population-based cohort studies using information from administrative records as outcomes, which may result in possible precursors of disorders going unreported and make accounting for confounders more limited. Overall, 23 out of 30 studies in the review had ADHD or ASD as outcome, compared to seven studies about other psychiatric or behavioral outcomes. In addition to the neurodevelopmental outcomes, antibiotics in the first 2 years of life were associated with major depressive disorder, but no sibling-controlled studies were done with this outcome. Thus, it seems that there is still less comprehensive knowledge on the internalizing and externalizing symptoms as outcomes in children. Prospective observational long-term studies could better address the potential causal roles of C-section and antibiotics on the full range of psychiatric outcomes while controlling for confounding factors.

One of the possible mechanisms explaining the association between early life exposures and later (neuro)psychiatric outcomes has been thought to be the gut-brain-axis^4^, with the GM of children born with C-section known to be altered compared to children born with vaginal delivery^37–39^. Dysbiosis of GM created by antibiotics in early life has been reported to persist for several months and even up to 2 years^12,40,41^. Stokholm et al. found that while GM composition was significantly altered between children born with vaginal delivery, emergency C-section and planned C-section, there were no colonization differences at 1 year after birth^37^. Our findings suggest that even though C-section and early antibiotics are known to alter infant GM, these exposures alone are unlikely to increase the risk for social-emotional problems in early childhood in a low-risk setting in a high-income country. In Finland, the C-section is often due to medical indication rather than a social reasons and decreasing antibiotics prescription rate renders more responsible antibiotics use in the nation^42,43^. Hence, the C-section and antibiotic use are often medically needed, and our null finding can be considered positive for the population health.

Given the considerable normative variability in questionnaire scores assessing social–emotional development among young children, detecting subtle associations within an unselected population may be challenging. Grisbrook et al. found no association between delivery mode and child behavior but discovered that C-section was indirectly associated with child internalizing and externalizing behaviors via maternal post-traumatic stress disorder and postpartum depression symptoms^44^. Hence, among certain risk groups, the association between early life exposures and later social-emotional problems might be detectable. In future studies, focusing on more clinically severe phenotypes (i.e., children who exhibit more pronounced symptoms than their peers and at an earlier age) and on specific risk groups, such as children exposed to maternal pre- or perinatal stress^45^ may provide a better understanding of this topic and direct the development of early interventions to support families.

There are several strengths in this study, including large sample size and use of national register data. It is one of the first studies to combine information on mode of delivery, prenatal, perinatal and early childhood antibiotics with child psychopathology. Our register data can be considered to reflect the use of outpatient antibiotics, since we used the data of not only prescribed, but purchased antibiotics. However, it is possible that not all purchased antibiotic courses have been taken. We did not consider any inpatient antibiotic treatments that might have been given in hospitals, apart from perinatal antibiotics given during delivery or up to 7 days postpartum. However, receiving inpatient (intravenous) antibiotic treatments is relatively rare in the general population of young children. Additionally, often, if a patient receives (non-oral) inpatient antibiotic courses, follow-up oral courses of antibiotics are often prescribed. Hence, we expect that the lack of information on inpatient antibiotic courses is unlikely to affect our conclusions. We only included child perinatal antibiotics, not mother’s. In Finland, it is clinical practice to give mother an antibiotic prophylaxis in all C-sections as well as some of the vaginal deliveries and this has not been included in the perinatal antibiotics in this study. Antibiotic prophylaxis is a single dose of cephalosporine or penicillin given intravenously less than an hour before delivery. This could affect child’s GM as an independent factor, although it seems that alterations in infant GM are more likely due to C-section itself than the prophylactic antibiotic dose^46^.

## Conclusions

Our results do not support the hypothesis that the mode of delivery or early life antibiotics would be associated with increased social and emotional problems in young children in the general population. Similar results in girls and boys indicate that these potential early life exposures do not associate with child outcomes in sex-specific manner. To the best of our knowledge, there are no other studies testing the combinatorial effects of C-section and early life antibiotics on child psychopathology in these age groups. As both C-section and antibiotic treatments are common and sometimes unavoidable interventions, our results might bring some relief for parents worried about possible long-term effects of these exposures.

## Ethics statement

The Joint Ethics Committee of South-Western Hospital District and the University of Turku gave positive statements to all sections of the study (VARHA/57/180/2011). This study has been conducted in according to the Declaration of Helsinki and European General Data Protection Regulations.

## Informed consent

Written informed consent was obtained from the study participants and parents gave consents on behalf of their children.

## Data sharing statement

Current EU and national legislation on personal data protection of sensitive data and the informed consents given by the study subjects do not permit open sharing of invidual-level data. The data can be shared as part of research collaboration with relevant Research Agreements in place. Investigators interested in research collaboration and obtaining access to the data can contact FinnBrain board.

## Funding and acknowledgements

We acknowledge Alice S. Carter for collaboration regarding BITSEA assessments. We thank the Turku University Foundation, the Research Council of Finland (#308176, #347640, #253270, #264363), Finnish State Grants for Clinical Research (ERVA), Signe and Ane Gyllenberg Foundation, Yrjö Jahnsson Foundation, Jalmari and Rauha Ahokas Foundation, Finnish Medical Foundation, Jane and Aatos Erkko Foundation, Stiftelsen Eschnerska Frilasarettet sr and Finnish Cultural Foundation for funding parts of this study.

