## Supplemental materials for "Early antibiotics, mode of delivery and social-emotional problems in childhood: The FinnBrain Birth Cohort Study"

Supplemental materials, Figure 1: *Potential confounders identified with a directed acyclic graph (DAG).*
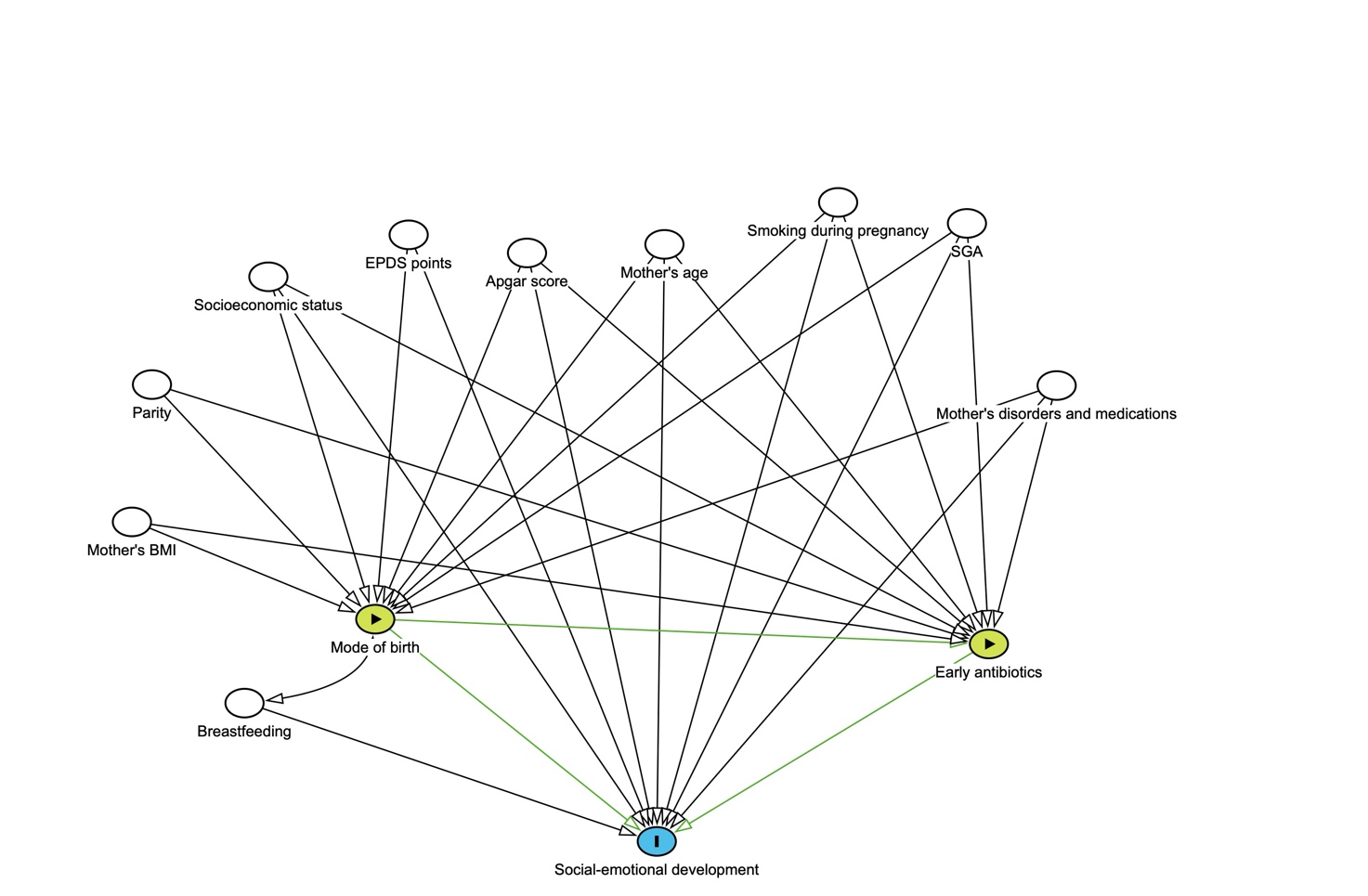

Supplemental materials, Table 1: *Attrition analysis was conducted to examine differences between participants who returned the questionnaires and those who did not.*

|  |  | Returned BITSEA | | | |
| --- | --- | --- | --- | --- | --- |
|  |  | No | Yes | p |  |
| Maternal age at delivery | mean | 29,64 | 31,11 | <0,001 | t-test |
| Maternal education | < 12 years | 731 | 385 | <0,001 | χ2 |
|  | 12-15 years | 473 | 395 |  |  |
|  | > 12 years | 421 | 563 |  |  |
| Birth mode | Vaginal | 1610 | 989 | 0,694 | χ2 |
|  | Vaginal, breach | 19 | 12 |  |  |
|  | Vacuum Extraction | 237 | 160 |  |  |
|  | Elective C-section | 124 | 82 |  |  |
|  | Urgent C-section | 200 | 139 |  |  |
|  | Emergency C-section | 25 | 22 |  |  |
| Parity | Multipara | 1173 | 654 | <0,001 | χ2 |
|  | Primipara | 1042 | 750 |  |  |
| Register data available | No | 14 | 4 | 0,148 | χ2 |
|  | Yes | 2201 | 1400 |  |  |
|  |  | **Returned SDQ** | | | |
|  |  | No | Yes | p |  |
| Maternal age at delivery | mean | 29,69 | 30,98 | <0,001 | t-test |
| Maternal education | < 12 years | 720 | 396 | <0,001 | χ2 |
|  | 12-15 years | 449 | 868 |  |  |
|  | > 12 years | 415 | 569 |  |  |
| Birth mode | Vaginal | 1594 | 1005 | 0,269 | χ2 |
|  | Vaginal, breach | 17 | 14 |  |  |
|  | Vacuum Extraction | 224 | 173 |  |  |
|  | Elective C-section | 119 | 87 |  |  |
|  | Urgent C-section | 192 | 147 |  |  |
|  | Emergency C-section | 28 | 19 |  |  |
| Parity | Multipara | 1156 | 671 | <0,001 | χ2 |
|  | Primipara | 1018 | 774 |  |  |
| Register data available | No | 14 | 4 | 0,124 | χ2 |
|  | Yes | 2160 | 1441 |  |  |
|  |  | **Returned either BITSEA or SDQ** | | | |
|  |  | No | Yes | p |  |
| Maternal age at delivery | mean | 29,52 | 30,93 | <0,001 | t-test |
| Maternal education | < 12 years | 605 | 511 | <0,001 | χ2 |
|  | 12-15 years | 363 | 505 |  |  |
|  | > 12 years | 310 | 674 |  |  |
| Birth mode | Vaginal | 1359 | 1240 | 0,457 | χ2 |
|  | Vaginal, breach | 14 | 17 |  |  |
|  | Vacuum Extraction | 192 | 205 |  |  |
|  | Elective C-section | 101 | 105 |  |  |
|  | Urgent C-section | 163 | 176 |  |  |
|  | Emergency C-section | 24 | 23 |  |  |
| Parity | Multipara | 991 | 836 | <0,001 | χ2 |
|  | Primipara | 862 | 930 |  |  |
| Register data available | No | 12 | 6 | 0,156 | χ2 |
|  | Yes | 1841 | 1760 |  |  |

Supplemental materials, Table 2: *BITSEA and SDQ scores in different groups.*

|  | Prenatal AB*:  no (n=1263) | yes (n=503) |  |  |
| --- | --- | --- | --- | --- |
| Bitsea Problem score |  |  |  |  |
| Mean (SD) | 7.45 (4.26) | 7.51 (4.37) |  |  |
| Median [Q1, Q3] | 7 [4, 10]] | 7 [4, 10] |  |  |
| Missing | 286 | 128 |  |  |
| SDQ sum score |  |  |  |  |
| Mean (SD) | 8.81 (4.87) | 9.04 (5.40) |  |  |
| Median [Q1, Q3] | 7 [5, 11] | 7 [5, 12] |  |  |
| Missing | 261 | 104 |  |  |
|  | Perinatal AB:  no (n=1575) | yes (n=191) |  |  |
| Bitsea Problem score |  |  |  |  |
| Mean (SD) | 7.48 (4.30) | 7.39 (4.20) |  |  |
| Median [Q1, Q3] | 7 [5, 10] | 7 [4, 9] |  |  |
| Missing | 374 | 40 |  |  |
| SDQ sum score |  |  |  |  |
| Mean (SD) | 8.79 (4.98) | 9.53 (5.37) |  |  |
| Median [Q1, Q3] | 8 [5, 11.75] | 9 [6, 12] |  |  |
| Missing | 331 | 34 |  |  |
|  | AB, first 2 yrs of life:  0 (n=612) | 1-3 (n=774) | 4 or more (n=380) |  |
| Bitsea Problem score |  |  |  |  |
| Mean (SD) | 7.37 (4.29) | 7.33 (4.30) | 7.93 (4.24) |  |
| Median [Q1, Q3] | 7 [4, 9] | 7 [4, 10] | 7 [5, 10] |  |
| Missing | 131 | 180 | 103 |  |
| SDQ sum score |  |  |  |  |
| Mean (SD) | 8.93 (5.10) | 8.71 (4.84) | 9.11 (5.29) |  |
| Median [Q1, Q3] | 8 [5, 12] | 8 [5, 11] | 8 [5, 12] |  |
| Missing | 131 | 160 | 74 |  |
|  | Delivery mode:  Assisted vaginal (n=222) | Emergency CS (n=199) | Planned CS (n=105) | Unassisted vaginal (n=1240) |
| Bitsea Problem score |  |  |  |  |
| Mean (SD) | 7.59 (3.92) | 7.14 (4.36) | 7.94 (4.93) | 7.46 (4.28) |
| Median [Q1, Q3] | 7 [[5, 10] | 6 [4, 9.08] | 7 [5, 10.83] | 7 [[4.35, 9.39] |
| Missing | 54 | 43 | 26 | 291 |
| SDQ sum score |  |  |  |  |
| Mean (SD) | 9.52 (5.16) | 8.99 (4.72) | 8.07 (5.19) | 8.80 (5.03) |
| Median [Q1, Q3] | 9 [6, 13] | 8 [6, 12] | 7 [[5, 11] | 8 [5, 11.5] |
| Missing | 39 | 37 | 21 | 268 |
|  | Delivery mode:  All vaginal (n=1462) | All C-sections (n=304) |  |  |
| Bitsea Problem score |  |  |  |  |
| Mean (SD) | 7.48 (4.23) | 7.41 (4.57) |  |  |
| Median [Q1, Q3] | 7 [5, 10] | 7 [4, 10] |  |  |
| Missing | 345 | 69 |  |  |
| SDQ sum score |  |  |  |  |
| Mean (SD) | 8.91 (5.06) | 8.68 (4.89) |  |  |
| Median [Q1, Q3] | 8 [5, 12] | 8 [5, 11] |  |  |
| Missing | 307 | 58 |  |  |
|  | AB (prenatal) + DM*  combination:  CS + no AB (n=214) | CS + AB (n=90) | VD + no AB (n=1049) | VD + AB(n=413) |
| Bitsea Problem score |  |  |  |  |
| Mean (SD) | 7.00 (4.09) | 8.43 (5.49) | 7.54 (4.29) | 7.32 (4.07) |
| Median [Q1, Q3] | 6 [4, 9] | 8 [4, 11] | 7 [5, 10] | 7 [4.5, 9] |
| Missing | 48 | 21 | 238 | 107 |
| SDQ sum score |  |  |  |  |
| Mean (SD) | 8.58 (4. 88) | 8.89 (4.95) | 8.85 (4.87) | 9.07 (5.51) |
| Median [Q1, Q3] | 8 [5, 12] | 8 [5.75, 10.25] | 8 [5.25, 11] | 8.88 [5, 12] |
| Missing | 44 | 14 | 217 | 90 |
|  | AB (perinatal) + DM  combination:  CS + no AB (n=255) | CS + AB (n=49) | VD + no AB (n=1320) | VD + AB (n=142) |
| Bitsea Problem score |  |  |  |  |
| Mean (SD) | 7.50 (4.51) | 6.97 (4.86) | 7.47 (4.26) | 7.54 (3.95) |
| Median [Q1, Q3] | 7 [4, 10] | 6 [4, 9.23] | 7 [5, 10] | 7 [5, 9] |
| Missing | 61 | 8 | 313 | 32 |
| SDQ sum score |  |  |  |  |
| Mean (SD) | 8.54 (4.86) | 9.38 (5.06) | 8.84 (5.00) | 9.57 (5.48) |
| Median [Q1, Q3] | 8 [5, 11] | 9 [5.75, 12] | 8 [5, 12] | 9 [6, 12] |
| Missing | 48 | 10 | 283 | 24 |
|  | AB (first 2 yrs of life) + DM combination:  CS + no AB (n=120) | CS + AB (n=184) | VD + no AB (n=492) | VD + AB (n=970) |
| Bitsea Problem score |  |  |  |  |
| Mean (SD) | 7.40 (4.6) | 7.42 (4.56) | 7.37 (4.22) | 7.54 (4.24) |
| Median [Q1, Q3] | 7 [4, 10.08] | 6.5 [4, 10] | 7 [4, 9] | 7 [5, 10] |
| Missing | 27 | 42 | 104 | 241 |
| SDQ sum score |  |  |  |  |
| Mean (SD) | 8.43 (4.76) | 8.83 (4.99) | 9.05 (5.18) | 8.85 (5.00) |
| Median [Q1, Q3] | 8 [5, 11] | 8 [5, 11] | 9 [5, 12] | 8 [5, 11] |
| Missing | 25 | 33 | 106 | 201 |

**AB = Recieved antibiotics*

*no AB = did not recieve antibiotics*

*CS = all caesarean secrions*

*VD= all vaginal deliveries*

*DM = Delivery mode*

*yrs= years*

Supplemental materials, Figure 2: *BITSEA scores in different groups.*

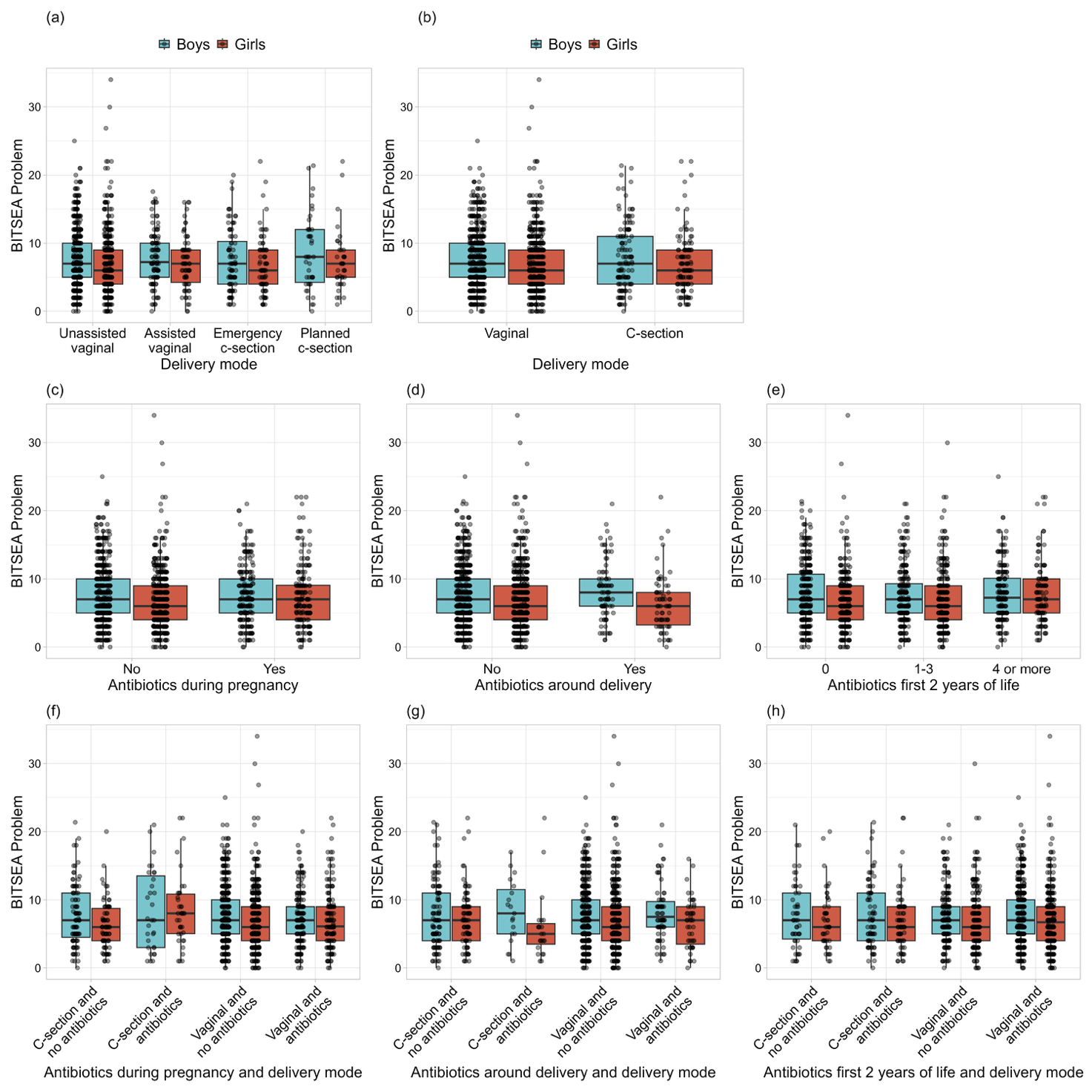

Supplemental materials, Figure 3: *SDQ scores in different groups*.

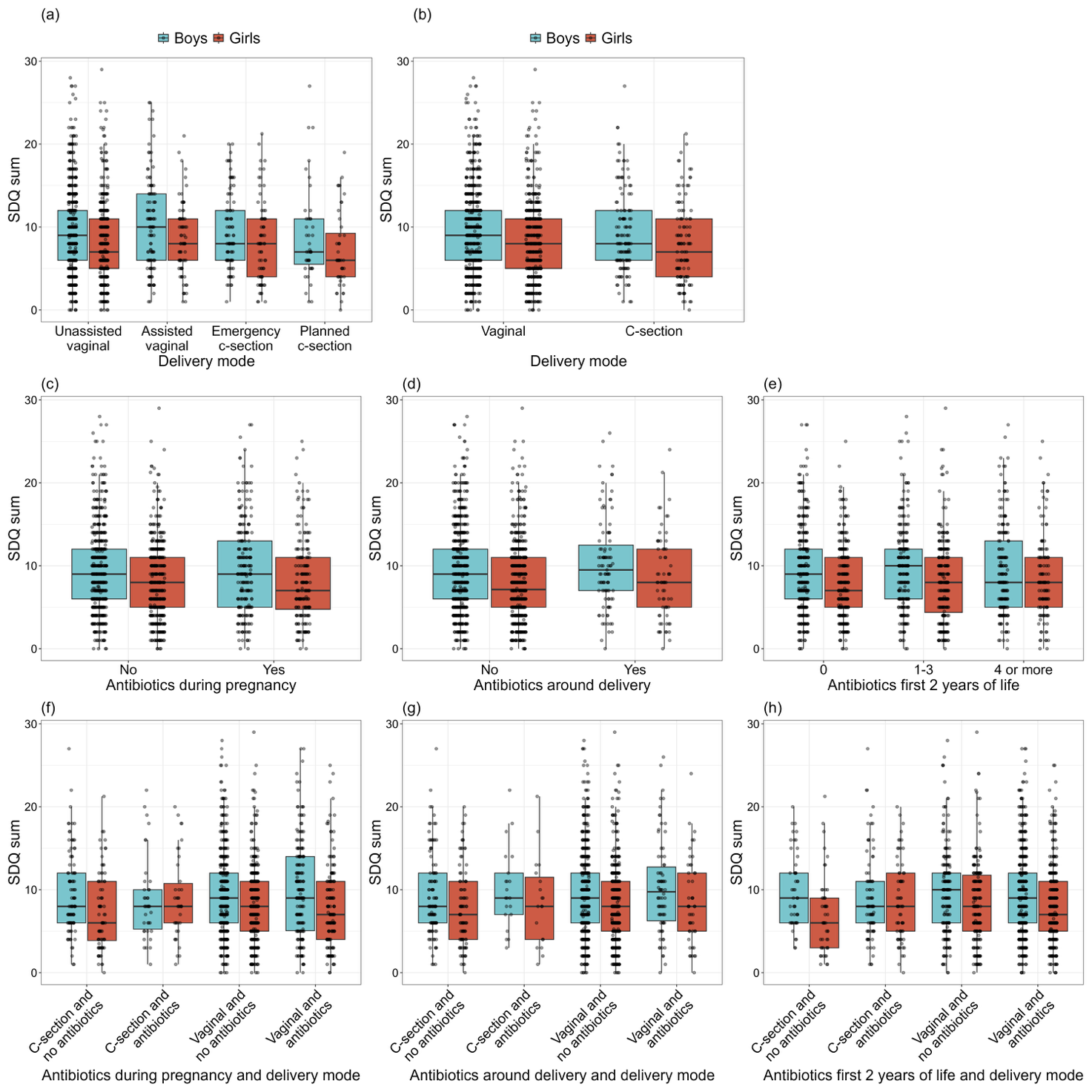
